# Causal relationships between gut microbiome and hundreds of age-related traits: evidence of a replicable effect on ApoM protein levels

**DOI:** 10.1101/2025.02.03.25321568

**Authors:** Federica Grosso, Daniela Zanetti, Serena Sanna

**Affiliations:** Institute for Genetic and Biomedical Research (IRGB) of the National Research Council (CNR), Monserrato (CA), Italy

**Keywords:** causal inference, aging, gut microbiome, inflammatory proteins, age-related macular degeneration

## Abstract

In the past 20 years, the involvement of gut microbiome in human health has received particular attention, but its contribution to age-related diseases remains unclear. To address this, we performed a comprehensive two-sample Mendelian randomization investigation, testing 55130 potential causal relationships between 37 traits representing gut microbiome composition and function and age-related phenotypes, including 1472 inflammatory and cardiometabolic circulating plasma proteins from UK Biobank Pharma Proteomic Project and 18 complex traits. A total of 91 causal relationships remained significant after multiple testing correction (false discovery rate p-value<0.05) and sensitivity analyses, notably two with the risk of developing age-related macular degeneration and 89 with plasma proteins. The link between purine nucleotides degradation II aerobic pathway and apolipoprotein M was further replicated using independent genome-wide association study data. Finally, by taking advantage of previously reported biological function of *Faecalibacterium prausnitzii* we found evidence of regulation of six proteins by its function as mucosal-A antigen utilization. These results support the role of gut microbiome as modulator of the inflammatory and cardiometabolic circuits, that may contribute to the onset of age-related diseases, albeit future studies are needed to investigate the underlying biological mechanisms.

## Introduction

Many human disorders, such as cancer, diabetes, neurodegeneration and cardiovascular diseases, are closely linked to the intricate and multifactorial process of aging, which involves physiological changes. To identify possible therapeutic targets for age-related disorders and to understand the mechanism of aging, it is essential to study changes within different body compartments during lifespan, such as those in the gut microbiome [1]. The human gastrointestinal tract (gut) hosts a vast microbial community of approximately 100 trillion microorganisms, which changes significantly throughout life. Furthermore, it is composed of a large number of immune cells that constantly communicate with the gut microbiota, an essential process for maintaining immune homeostasis. Disruption of this interaction can lead to dysbiosis, which can contribute to the development of autoimmune diseases, inflammatory diseases, cardiovascular diseases, susceptibility to infections, and other health problems [2], [3]. Therefore, changes in the gut microbiota influence various aspects of both gut and systemic immune and inflammatory responses, suggesting a link to the age-related decline in cardiovascular and immune function, often referred to as immune aging, immunosenescence or inflammaging [3].

During aging, significant changes occur in the gut microbiome. For example, opportunistic pathogens such as *Enterobacteria*, which can induce intestinal inflammation, increase, while by contrast beneficial commensals such as *Bacteroides*, *Bifidobacteria* and *Lactobacilli* decrease [3]. These changes in microbial composition underline the importance of studying the microbiota throughout life to better understand its role in aging and associated diseases. However, it remains uncertain whether changes in the microbiota drive these conditions or whether these or the related therapies impact the microbiota. Given this ambiguity, we investigated the causal relationships between the gut microbiome and age-related complex traits, along with associated immune-related and cardiometabolic circulating proteins.

To investigate potential causal links between the gut microbiome and age-related traits and proteins, the two samples Mendelian randomization (MR) technique was employed, a causal inference approach that uses genome-wide association study (GWAS) data to infer causal relationships while controlling for confounding factors and reverse causality. Other studies [4], [5], [6], [7], [8], [9], [10] have used MR to assess the causal relationships between gut microbiome and specific age-related traits. For example, Mao et al. [6] investigated its causality with age-related macular degeneration; Chen et al. [7], [10] explored links with longevity traits; and Bo et al. [8] focused specifically on frailty. In our work, we opted for a comprehensive analysis with an extensive number of age-related outcomes. Furthermore, we emphasized the importance of a rigorous approach to MR analyses. This involves i) employing GWAS carried out on large data set to ensure sufficient statistical power, ii) the use of stringent parameters to ensure validity of MR assumptions such as the independence and strong association of the instrumental variables (IVs), iii) performing multiple testing correction and sensitivity analysis, and iv) exclusion of reverse causality bias via bidirectional MR. Moreover, our study followed the STROBE-MR (Strengthening the Reporting of Observational Studies in Epidemiology using Mendelian Randomization) checklist [11] and guidelines from Burgess et al. [12].

Unlike previous studies, we performed replication analyses for the significant results using independent GWAS datasets, a fundamental step that has often been overlooked. Replication [13] is one way to prevent false positive results from being spread [14], [15]. This approach strengthens the robustness and reliability of our findings, making them a more solid reference point for understanding the causal relationships between the gut microbiome and aging-related phenotypes.

## Results

A two-sample MR was performed to investigate the causal relationships between the gut microbiome (exposure) and selected age-related phenotypes (outcome). As exposure we considered 37 microbiome features, with at least one variant associated at genome-wide significant level with the trait (**Supplementary Table 1a**). All the exposures showed an F-statistic > 10, indicating sufficient instrument strength and reducing the risk of weak instrument bias (**Supplementary Table 2a**). As outcomes we selected 1490 age-related phenotypes including diseases and quantitative traits **(Table 1)**. We searched for publicly available GWASs related to the most common age-related diseases, excluding cancer and neurological disorders, and for traits related to overall aging (like lifespan and longevity). We identified 18 age-related trait GWASs that included at least 20,000 European ancestry individuals and for which the necessary information to perform MR analysis (SNP chromosome and position, effect allele, other allele, beta, standard error, p-value) was available. We also included 1472 inflammatory and cardiometabolic proteins, selected from the study of Sun et al., 2023 [16].

**Table 1.** Age-related outcomes selected for the MR analysis. This table shows all the GWAS summary statistics used as outcomes in the MR analysis and the reason for the choice of these specific GWAS. GWAS: genome-wide association study, MR: mendelian randomization.

| OUTCOME NAME | DATA ARTICLE | SAMPLE SIZE | TYPE OF TRAIT | CHOICE OF THE TRAIT |
| --- | --- | --- | --- | --- |
| Age-related macular degeneration | Jiang et al., 2021 (PMID: 34737426) | 456348: 1295 cases and 455053 controls | Binary (ICD-9-CM: 362.29, based on hospital record) | European ancestry with higher sample size |
| Cardiovascular ageing | Shah M. et al., 2023 (PMID: 37604819) | 29506 | Continuous | European ancestry with higher sample size |
| Coronary artery disease | van der Harst P et al., 2017 (PMID: 29212778) | 296525: 34541 cases and 261984 controls | Binary | European ancestry with higher sample size |
| Frailty | Atkins JL et al., 2021 (PMID: 34431594) | 175226 | Continuous | European ancestry with higher sample size |
| Health span | Zenin A et al., 2019 (PMID: 30729179) | 300447 | Continuous | European ancestry with higher sample size |
| Ischemic stroke | Malik R. et al, 2018 (PMID: 29531354) | 440,328: 34,217 cases and 406,111 controls | Binary | European ancestry with higher sample size |
| Lifespan | Timmers et al., 2019 (PMID: 30642433) | 1012240 | Continuous | European ancestry with higher sample size |
| Longevity 90th percentile | Deelen J et al., 2019 (PMID: 31413261) | 36745: 11262 cases and 25483 controls | Binary | European ancestry with higher sample size |
| Longevity 99th percentile | Deelen J et al., 2019 (PMID: 31413261) | 28967: 3484 cases and 25483 controls | Binary | European ancestry with higher sample size |
| Osteoporosis | Dönertaş HM et al., 2021 (PMID: 33959723) | 484598: 7751 cases and 476847 controls | Binary | European ancestry with higher sample size |
| Parental longevity (combined parental attained age) | Pilling LC et al., 2017 (PMID: 29227965) | 389166 | Continuous | European ancestry with higher sample size |
| Parental longevity (both parents in top 10%) | Pilling LC et al., 2017 (PMID: 29227965) | 86949 | Continuous | European ancestry with higher sample size |
| Parental longevity (mother's age at death) | Pilling LC et al., 2017 (PMID: 29227965) | 246941 | Continuous | European ancestry with higher sample size |
| Parental longevity (mother's attained age) | Pilling LC et al., 2017 (PMID: 29227965) | 412937 | Continuous | European ancestry with higher sample size |
| Parental longevity (father's age at death) | Pilling LC et al., 2017 (PMID: 29227965) | 317652 | Continuous | European ancestry with higher sample size |
| Parental longevity (father's attained age) | Pilling LC et al., 2017 (PMID: 29227965) | 415311 | Continuous | European ancestry with higher sample size |
| Parental longevity (combined parental age at death) | Pilling LC et al., 2017 (PMID: 29227965) | 208118 | Continuous | European ancestry with higher sample size |
| Type 2 diabetes | Loh PR et al., 2018 (PMID: 29892013) | 468298 | Continuous | European ancestry with higher sample size |
| Circulating inflammatory and cardiometabolic proteins | Sun et al., 2023 (PMID: 37794186) | 54219 | Continuous (OLINK) | Most extensive recent panel of circulating inflammatory and cardiovascular proteins |

Out of the 55130 MR tests performed, 91 causal relationships remained significant after multiple-testing correction (FDR p-value<0.05). The results highlighted a causal link between the gut microbiome and age-related macular degeneration (AMD) and levels of 62 distinct circulating protein levels (**Supplementary Table 2**).

### Age-related macular degeneration

AMD GWAS was collected from the study of Jiang et al. [17], which analyzed data of 1,295 cases and 455,053 controls from UK Biobank [18]. We found that a genetic predisposition to higher levels of bacteria of the order of *Coriobacteriales* or of family *Coriobacteriaceae* increases the risk of developing AMD disease (*β*_*c*_ = 0.61, corresponding to an OR=1.84, *p*_*BH*_ = 0.047). All the sensitivity analyses **(Table 2)** and graphical inspections **(Supplementary Figures 2 and 3**) confirmed this result, further supported by the absence of reverse causality (all *p* > 0.87).

**Table 2.** Significant causal relationships between gut microbiome features and age-related macular degeneration. This table shows significant results of the MR analysis between gut microbiome and age-related macular degeneration. For each exposure-outcome pair, we show the causal estimate *β*_*c*_ (corresponding effect on disease risk liability for 1 standard deviation unit increase on the gut microbiome trait), the p-value obtained with IVW method, after FDR multiple testing correction, and the p-value from all sensitivity analyses (weighted median, MR-PRESSO as pleiotropy aware MR-tests, tests of pleiotropy (Egger intercept) and heterogeneity (Cochran’s Q), and the reverse causality). The last column indicates the lowest p-value of the replicated causal relationship using independent datasets. (AMD: age-related macular degeneration, FDR: false discovery rate, IVW: inverse variance weighted, MR: Mendelian randomization, WM: weighted median)

| EXPOSURE | OUTCOME | $\beta_c$ | IVW post-FDR correction pval | WM pval | MR-PRESSO pval | Pleiotropy pval | Heterogeneity pval | Reverse causality pval | Lowest replication pval |
| --- | --- | --- | --- | --- | --- | --- | --- | --- | --- |
| *Coriobacteriales* | AMD | 0.61 | 0.047 | 0.007 | 0.009 | 0.44 | 0.75 | 0.98 | 0.234 |
| *Coriobacteriaceae* | AMD | 0.61 | 0.047 | 0.007 | 0.009 | 0.44 | 0.75 | 0.98 | 0.234 |

We tested the replicability of these causal relationships using three more GWAS data of AMD **(Supplementary Table 1c)**. Albeit the definition of AMD was different, their large sample size provided sufficient power for replication **(Supplementary Figure 5)**, i.e. p-value<0.05 and same direction of the effect as in the main MR analysis. Despite the sufficient power, none of these three GWASs led to significant results **(Supplementary Figure 4)**.

### Cardiometabolic and inflammatory proteins

The GWAS summary statistics for circulating inflammatory and cardiometabolic proteins refer to the study of Sun et al., 2023 [16], which analyzed a total of 2923 proteins in 54219 UK Biobank participants **(Supplementary Table 1b)**. Among all these proteins, we analyzed as outcomes a total of 1472 proteins from the inflammatory and cardiometabolic panels, markers of interest for our study. Out of all the 54464 MR analyses carried out with the 37 microbiome features, 89 showed significant MR results after FDR correction (adjusted p<0.05) and all sensitivity analyses.

Interestingly, out of these 89 total causal relationships detected, 57 were with proteins listed in the inflammatory panel (36 distinct proteins) and 32 with proteins from the cardiometabolic panel (26 distinct proteins) (**Figure 2**).

**Figure 1.**
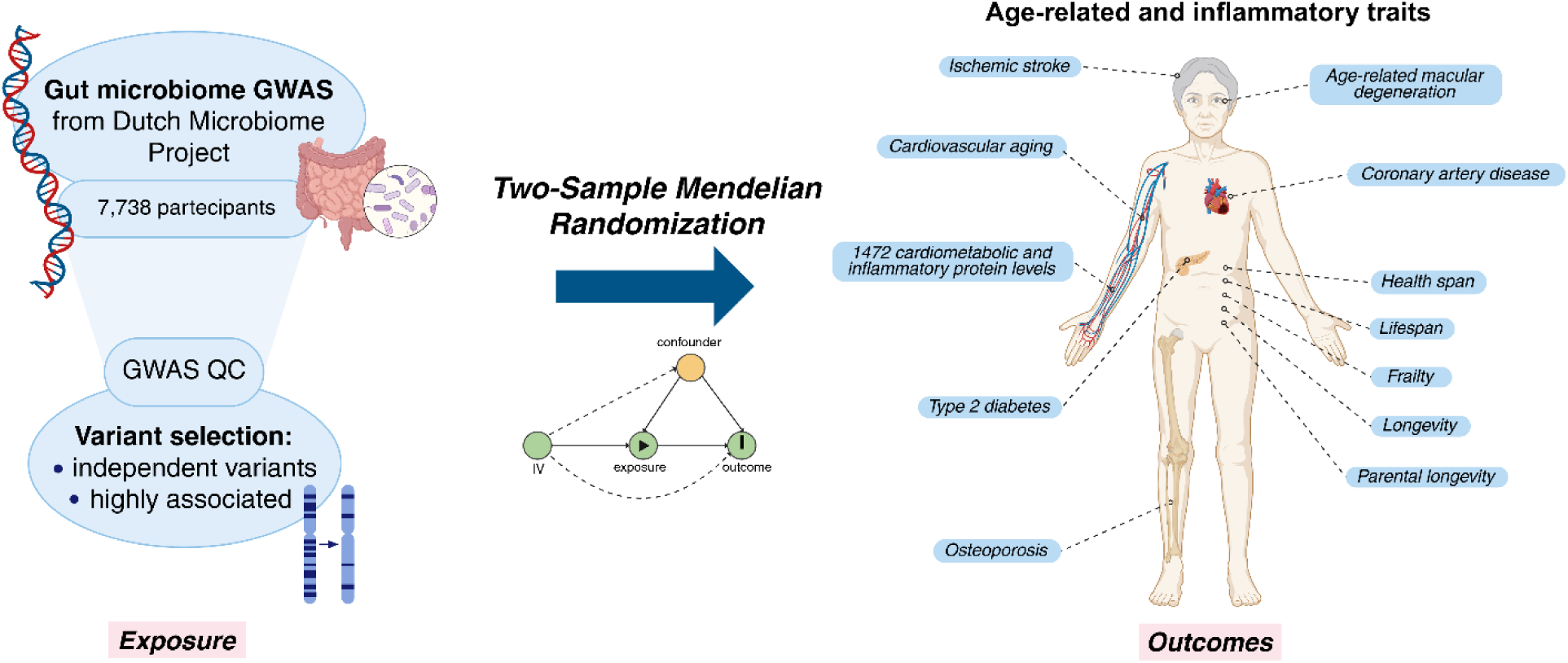
Graphical representation of the study. Created in BioRender. Sanna, S. (2025) https://BioRender.com/a45o861

**Figure 2.**
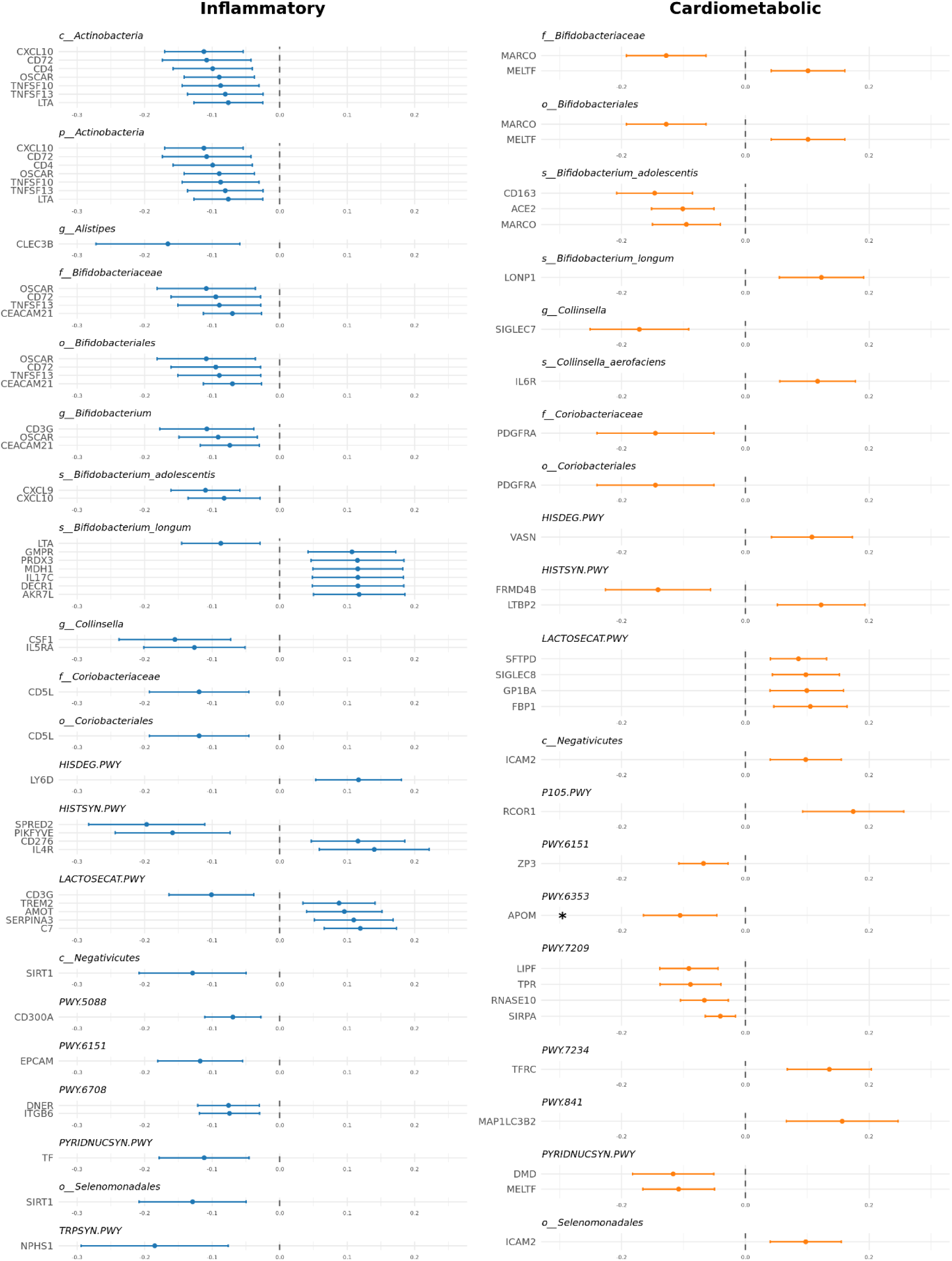
Significant causal relationships between gut microbiome and inflammatory and cardiometabolic proteins. Forest plot showing causal estimates for the 89 significant causal relationships identified.

We replicated significant causal relationships for proteins for which GWASs were available in other studies. Specifically, we used three key datasets: Zhao et al. [19], Sun et al., 2018 [20], and Folkersen et al. [21] (**Supplementary Table 1c**). A relationship was considered replicated if the IVW p-value was less than 0.05 and the direction of the effect was consistent with our main analysis. With these criteria, we found that the causal relationship between the purine nucleotides degradation II aerobic pathway and apolipoprotein M (ApoM) was replicated using the ApoM GWAS from Sun et al., 2018 [20] (GCST90240318) (**Figure 3**). In particular, in both analyses an increase in pathways abundance was causally linked with a decrease in ApoM circulating levels (main analysis: *β*_*c*_ = −0.11, *p*_*BH*_ = 0.02; replication analysis: *β*_*c*_ = −0.22, *p* = 0.01). All other results are detailed in the **Supplementary Table 2k**.

**Figure 3.**
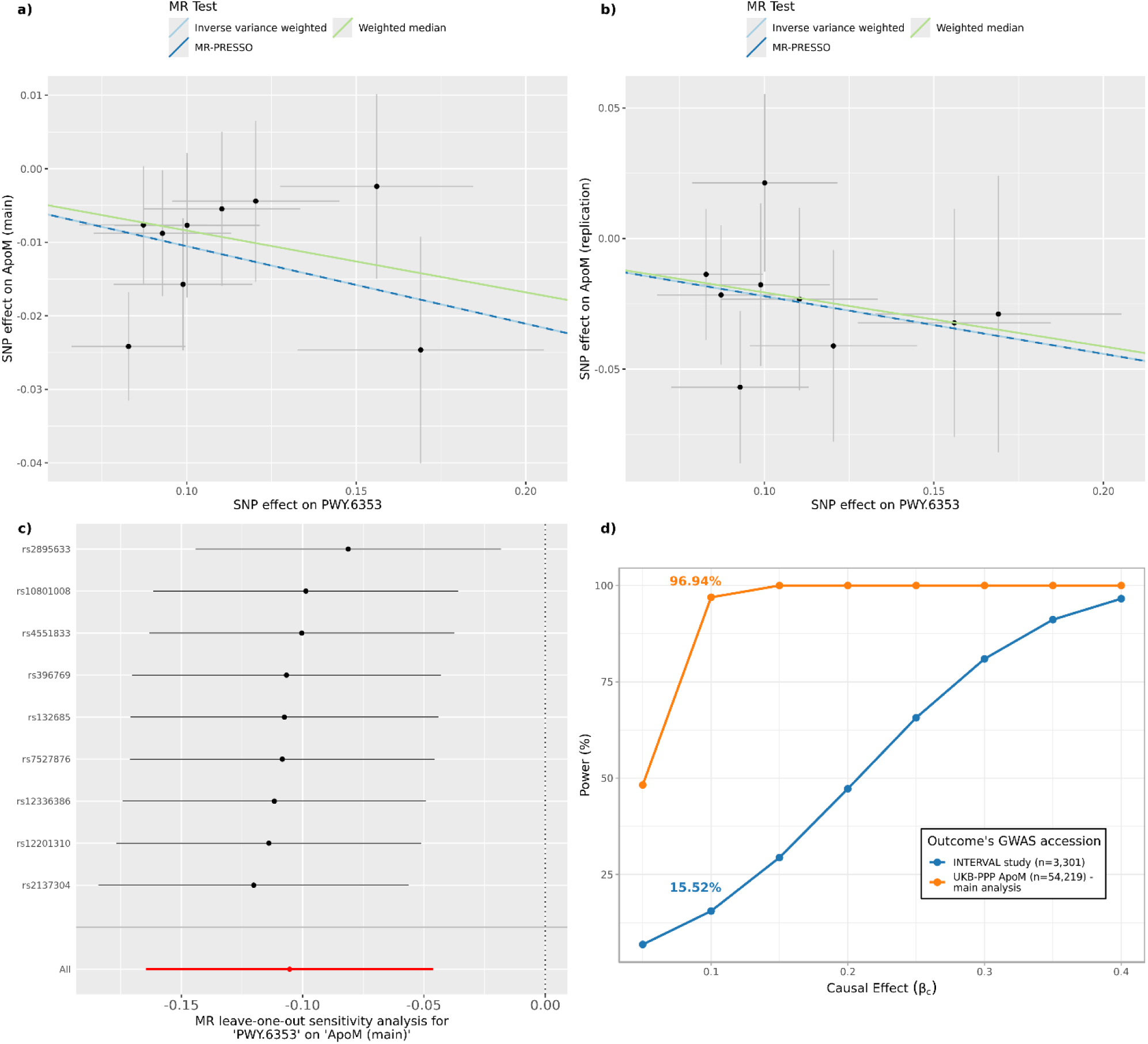
The causal relationship between purine nucleotides degradation pathway and ApoM levels. The **panel a)** shows the results of the main MR analysis between purine nucleotides degradation II aerobic pathway and ApoM protein. In the scatter plot, each dot is an IV and the x and y-axis represents the association coefficients with the exposure and outcome, respectively. The three lines represent the results of the three different MR tests, with the slope of the lines being equal to causal estimates of each test. The **panel b)** follows the same definition of panel a) but refers to results obtained using the GWAS data of the ApoM protein from Sun et al., 2018 (GCST90240318) as outcome. In the **panel c)** the causal estimates from the leave-one-out analyses are shown and compared with the causal estimate from main analysis (red line). In the **panel d)** we show the post-hoc power estimates at varying causal effect sizes for different studies of ApoM as outcome. The causal effect identified in the main analysis (*β*_*c*_ = -0.11) and corresponding power are highlighted in the curves. (ApoM: Apolipoprotein M; GWAS: genome-wide association study, IV: instrumental variable, MR: Mendelian randomization, PWY.6353: purine nucleotides degradation II aerobic pathway, SNP: single nucleotide polymorphism)

Of note, unlike the UK Biobank Plasma Proteomic Project (UKB-PPP) proteins used in Sun et al., 2023 [16], measured using the OLINK platform, the INTERVAL study (Sun et al., 2018 [20]) employed the SOMAscan assay for protein quantification. Additionally, the INTERVAL study has a much smaller sample size compared to the UK Biobank (3622 vs. 54219 participants). In contrast, the other two studies – Folkersen et al. [21], and Zhao et al. [19] – also used OLINK for protein measurements albeit with a slightly different methodology, and have sample sizes of 21758 and 14824 participants, respectively.

### Further investigation of GalNAc-linked utilization

We further investigated the underlying biological meaning of the significant causal relationships between the lactose-galactose degradation I pathway (LACTOSECAT) pathway, and plasma protein levels, being this pathway strongly associated with a genetic variant in the *ABO* gene [22]. We therefore aimed to investigate whether the causal relationships observed in our study might be connected to the utilization of N-acetylgalactosamine (GalNAc) in blood type A individuals, since a recent study from our collaborators [23] showed that strains of *ABO*-associated species, such as strains of *Faecalibacterium prausnitzii*, can utilize this sugar from individuals who secrete the mucosal A-antigen.

We conducted MR analysis using as exposures host genetic variants associated with the deletion SV (dSV) region (577-579 kb) present in strains of *F. prausnitzii* known to be active in degradation of secreted mucosal A-antigens (**Supplementary Table 1d**), and proteins and LACTOSECAT pathway as outcomes. First, we corroborated the existing biological evidence with MR, and showed that predisposition to higher load of *F. prausnitzii* SVs (thus higher GalNAc utilization activity) is causally linked with higher levels of the lactose-galactose degradation I pathway, supporting our hypothesis that the activity of this microbial pathway is related to mucosal-A antigen utilization (IVW *p* < 0.05 for all SVs) (**Supplementary Table 2r**). We also detected a causal link between *F. prausnitzii* SV and 2 cardiometabolic and 4 inflammatory protein levels in blood. Specifically, we found that higher load of *F. prausnitzii* SVs and consequently higher mucosal-A antigen utilization, was causally linked with an increase in circulating levels of Pulmonary surfactant-associated protein D (SFTPD), Sialic acid-binding Ig-like lectin 8 (SIGLEC8), Triggering receptor expressed on myeloid cells 2 (TREM2), Alpha-1-antichymotrypsin (SERPINA3) and Complement component C7 (C7), and with decreasing levels of T-cell surface glycoprotein CD3 gamma chain (CD3G) (all IVW p <0.05). Furthermore, the direction of the effect of the MR analysis between *F. prausnitzii* SV and proteins is consistent with the direction of the effect of the MR analysis detected between LACTOSECAT pathway and proteins (**Figure 4**).

**Figure 4.**
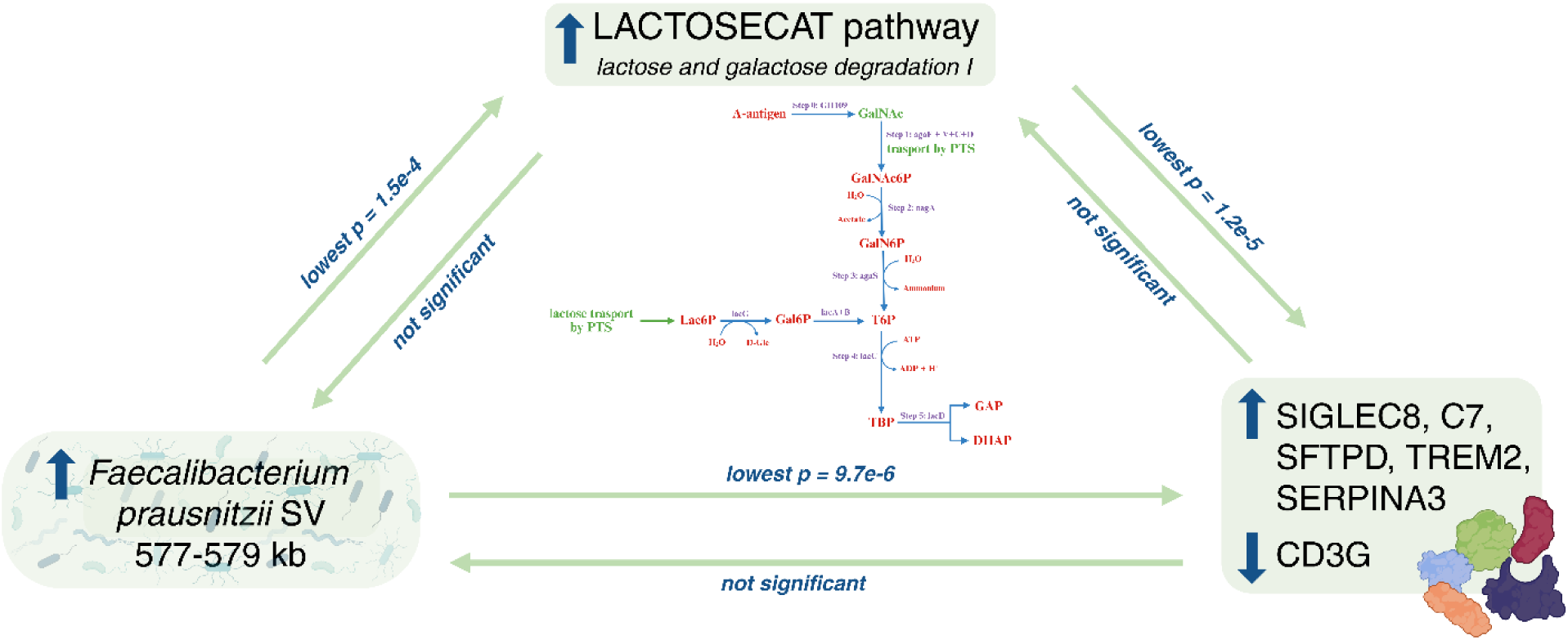
Directed causal relationships involving the abundance of *Faecalibacterium prausnitzii structural variant 577_579*, the LACTOSECAT pathway, and plasma proteins. The figure illustrates the causal relationship between the abundance of *Faecalibacterium prausnitzii* structural variant 577_579 and LACTOSECAT pathway, between LACTOSECAT pathway and six plasma proteins and the one between *Faecalibacterium prausnitzii* and the same six plasma proteins. It also shows the lactose and galactose degradation I pathway (horizontal) along with the GalNAc degradation pathway (vertical), with annotated steps detailing the processes involved in GalNAc degradation, as described in Zhernakova et al. (2024). Created in BioRender. Sanna, S. (2025) https://BioRender.com/o9k1q5m (ApoM: Apolipoprotein M; DHAP: glycerone phosphate; GalNAc: N-acetylgalactosamine; Gal6P: D-galactopyranose 6-phosphate; GalNAc6P: N-acetyl-D-glucosamine-6-phosphate; GalN6P: N-Acetylgalactosamine-6-phosphate; GAP: D-glyceraldehyde 3-phosphate; LACTOSECAT pathway: lactose and galactose degradation pathway; Lac6P: lactose 6’-phosphate; T6P: D-tagatofuranose-6-phosphate; TBP: D-tagatofuranose 1,6-biphosphate)

## Discussion

In this study, we investigated the causal effect of variation in gut microbiome composition and function on age-related traits, using the Mendelian randomization approach. Among all the investigated outcomes, the analyses highlighted a potential causal effect on AMD, and on 36 inflammatory proteins and 26 cardiometabolic circulating plasma protein levels.

A strong and replicable causal relationship highlighted by our study is the one between purine nucleotides degradation II (aerobic) pathway and apolipoprotein M protein in plasma. This pathway leads to the stepwise breakdown of purine nucleotides into urate, which in humans is the final product due to the absence of the enzyme uricase. Urate is mostly reabsorbed by the kidneys, with only ∼10% excreted [24]. Its accumulation has been linked to cardiovascular risk - hyperuricemia is associated with higher incidence of cardiovascular events [25], [26]. Our findings suggest that increased purine degradation may contribute to cardiovascular risk through two converging mechanisms: elevated urate levels and reduced ApoM concentrations. ApoM, mainly produced in the liver and kidneys, is essential for HDL formation and function, promoting cholesterol efflux and exerting anti-inflammatory and atheroprotective effects. Its location in the Major Histocompatibility Complex (MHC) class III region also links it to immune and inflammatory processes [27]. An increase in urate levels may be a risk factor for endothelial dysfunction [28], a process that, particularly in older individuals who are more vulnerable to chronic inflammation, can lead to downregulation of ApoM expression. This may contribute to the development of atherosclerosis and subsequent vascular events [29].

Another interesting result is the identification of causal relationships between the LACTOSECAT pathway and six circulating protein levels, which we showed to be attributable to GalNAc utilization of secreted mucosal A antigen in blood type A individuals. Using MR and the existing knowledge of host-interaction function of *F. prausnitzii* strains, we confirmed that higher abundance of LACTOSECAT pathway implicates higher GalNAc utilization in blood type A individuals, and this in turn increases or decreases the levels of these proteins. Specifically, GalNAc utilization in blood type A individuals leads to higher activity of the pathway, and in turn to increased levels of SIGLEC8, SFTPD, TREM2, SERPINA3, and C7. Whereas CD3G levels decrease with both higher *F. prausnitzii* and increased LACTOSECAT pathway activity. Based on these results, we speculate that differences in disease risk among blood type groups for age-related conditions on which these proteins are potentially involved may be different due to a modulation of gut microbiome on host protein levels. In particular some studies demonstrated that TREM2 is protective for atherosclerosis [30], while the increase of the presence of some other proteins, like SERPINA3 and C7 led to an increase of cardiac mortality and coronary artery disease respectively [31], [32].

We acknowledge that there is an extensive growth in literature on the associations between gut microbiome and the onset of many chronic and age-related diseases. However, reliability and reproducibility of most of these findings are limited by several pitfalls in the statistical protocol used, such as a lenient selection of IVs, being limited to one single MR test and not performing sensitivity analyses, and the lack of correction for multiple testing. Furthermore, very few studies have aimed to replicate their findings in independent datasets, leaving their results inconclusive.

Chen et al. [7], [10], for example, investigated the presence of potential causal relationships between gut microbiome and five longevity traits – frailty, health span, lifespan, longevity, parental longevity – using the same GWAS data that we have used in our study. Among their exposures there were traits derived from the GWAS carried out on the Dutch Microbiome Project, which we also used in our study. They identified several causal relationships that we were unable to reproduce. The primary differences between their study and ours lie in the methodology, particularly in the threshold used for selecting IVs. Indeed, to avoid overestimating the causal effect, we only focused on microbiome traits with at least one genome-wide significant hit and opted for a more stringent association p-value of 5 ⋅ 10^−6^, instead of 1 ⋅ 10^−5^. Furthermore, we did not base our significance on nominal p-values but rather employed multiple testing correction. This approach reduces the number of significant findings but strengthens the robustness and reliability of our results. Out of the 168 exposure-outcome significant pairs reported by Chen et al. [7], [10], 12 would survive our pipeline for selection of exposure and only 3 were nominal significant for the IVW test. However, none were significant after adjusting for multiple testing (all *p*_*BH*_>0.16). Additionally, none would pass the sensitivity analyses therefore they would not be considered significant even without applying FDR (**Supplementary Table 3**).

Of note, another study (Mao et al. [6]) has specifically investigated the causal link between gut microbiome and AMD, but the two GWAS used for analyses were different from those used by us. As exposure (microbiome GWASs) they used data from the MiBioGen consortium [23], and as outcome (AMD GWAS) data were derived from FinnGen biobank analysis (round 5), which includes 3763 cases and 205359 controls. Using a *p* < 1 ⋅ 10^−5^as significant threshold to select IV, 6 microbiome features (out of 211 tested) were found to be causally linked to AMD at p < 0.05 (smallest p=0.005). None of these features include species of the order of *Coriobacteriales*. These results, however, were not supported by a correction for multiple testing, therefore they could not be considered statistically significant. In addition, the other MR methods they implemented were not taken into account; for example, the p-values from MR-PRESSO were never significant (all p>0.05). In contrast, the causal relationship we detected with AMD was robust to FDR correction and to all sensitivity analysis we opted for. It must be noted however that despite we have used a stringent and rigorous pipeline, we were unable to replicate the causal relationship using different datasets.

Our findings highlight the importance of a rigorous methodological approach in the application of MR to study the causal relationships between the gut microbiome and age-related phenotypes. A strict observance to MR guidelines [12], [33] and STROBE-MR [11] is essential to ensure the integrity and reliability of the results and a proper evaluation of finding, concept that extends beyond the specific exposure and outcome we selected. Several findings from previous studies [6], [7], [10] that did not follow the guidelines were in fact not supported by our analyses. Our study also focus attention on the need for replication of the results in independent GWAS datasets, rather than assuming that an initial statistical significance can be interpreted as definitive proof of causality. This concept is essential to avoid misinterpretation and to progress towards the discovery of effective therapeutic interventions. The range of applications of MR and related methods for understanding causal mechanisms has in fact expanded rapidly over the past 20 years, coupled with the increasing number of public GWAS studies, and therefore the adherence to a rigorous pipeline is mandatory.

We also acknowledge the study limitations. First, the genetic of microbiome traits has a limited impact on explaining the total variation, therefore only one IV could be selected at 5 ⋅ 10^−8^for each trait, significantly impacting power of MR. We mitigated this limitation by using a lower threshold of 5 ⋅ 10^−6^ to detect more IVs and rigorously assessed the coherence of causal estimates from each IVs using several sensitivity analysis methods. Secondly, we acknowledge that genetics of gut microbiome traits can be influenced by other factors, including diet and lifestyle factors which may confound MR analyses by deviating from the independence assumption. While we have no data to explore this confounding, we attempted to reduce its impact by considering only those microbiome traits with strong genetic link (at least one genome-wide significant hit), most of which are known to consistently replicate in other cohorts regardless of diet [34]. It must be noted that some of these effects may be even stronger depending on diet. For example, the association between ABO genetic variants with bacterial species and pathways, many of which show significant causal relationships in our analyses, is enhanced by fiber intake [35]. Third, we recognize the lack of replication of some results in independent cohorts. Furthermore, we are aware that differences in data collection methods and characteristics of the cohorts analyzed may have introduced sources of bias, which may have prevented the detection of weaker but still relevant relationships. For instance, in the case of AMD we identified three independent GWASs for the disease, each employing different case definitions. The primary AMD dataset considered AMD as a binary trait coded as 362.29 in the ICD-9-CM (**Supplementary Table 1b**), which differs from the definitions used in the other three datasets employed for replication (**Supplementary Table 1c**). Thus, despite the higher statistical power in the replication datasets (**Supplementary Figure 5**), inconsistent phenotypic definitions may have confounded replication results. Similarly, in the case of circulating inflammatory and cardiometabolic proteins, all replication datasets had lower sample sizes and thus lower power compared with the main analyses.

While our analyses are robust and the results compelling, we were unable to elucidate the underlying biological mechanisms driving the observed causal relationships. Therefore, any clinical application aimed at microbiome modulation remains premature.

In conclusion, we investigated the causal relationships between the gut microbiome and age-related phenotypes using the MR approach and ensured the integrity and reliability of the results by a strict adherence to MR guidelines. Our results support a causal role of gut microbiome in age-related macular degeneration and in both upregulating and downregulating the expression of 36 inflammatory and 26 cardiometabolic protein levels, some of which occur via mucosal-A antigen utilization. Particularly robust was the causal link between a microbial purine nucleotides degradation II aerobic pathway and and levels of the protein ApoM, which was successfully replicated in an independent cohort. While these links are particularly intriguing, we acknowledge that future studies are needed to investigate the underlying biological mechanisms and to further confirm our evidence.

## Materials and Methods

### Exposure and outcome selection

Two-sample MR was performed to investigate the causal relationship between the gut microbiome (exposure) and selected age-related phenotypes (outcome) (**Figure 1**). As exposure data we used genome-wide association study (GWAS) summary statistics collected from a previous study by Lopera-Maya et al. [22]. Lopera-Maya et al. used shotgun genome sequencing on fecal samples from 7738 individuals participating in the Dutch Microbiome Project [36] to derive quantitative information on 207 taxa and 205 pathways. By analyzing these 412 microbiome variables with GWAS method, they identified significant genetic associations (*p* < 5 ⋅ 10^−8^) for 37 of these: 18 taxa and 19 pathways; these 37 were considered as exposure for our study. The relevant GWAS summary statistics can be accessed through the GWAS Catalog (https://www.ebi.ac.uk/gwas/home) [37] **(Supplementary Table 1a).**

The outcomes of interest included 18 age-related phenotypes, both diseases and quantitative traits **(Table 1)**, of which summarized GWAS results were also collected on GWAS Catalog [37] and, for “lifespan”, from the Edinburgh DataShare repository (https://datashare.ed.ac.uk/handle/10283/3209). Additionally, we included 1472 circulating proteins from inflammatory and cardiometabolic panels of UK Biobank Plasma Proteomic Project (UKB-PPP), for which the GWAS summary statistics were reported in the study by Sun et al., 2023 [16]. These outcomes were selected for their relevance to aging and cardiometabolic health, as they comprise key traits related to longevity, frailty, cardiovascular and metabolic disorders, as well as systemic inflammation. Furthermore, these GWASs were selected because they were carried out in very large cohorts of European ancestry (28967<N<1012240), (**Table 1**), so they provided sufficient statistical power for analyses.

### GWAS quality control and selection of instrumental variables

We performed a quality control (QC) screening for each of the downloaded GWAS files (37 for the microbiome and 1490 for the outcomes). In particular, we confirmed that all studies were aligned to build 37 (genome assembly GRCh37/hg19) or otherwise to a compatible assembly such as hg38 (cardiovascular aging and circulating protein levels) and assigned missing rsIDs to genetic variants when needed. Furthermore, we removed variants in the exposure if their effect size was very large and likely unreliable (+/-4 standard deviation units).

After QC, we selected genetic variants to be used as IVs for MR analyses. Since MR assumptions require independent (not in linkage disequilibrium) and highly associated IVs, we selected genome-wide significant (*p* < 5 ⋅ 10^−6^) independent hits, after linkage disequilibrium clumping using a window of 10 *Mb* and *R*^2^ cutoff = 0.001.

To avoid the weak instrument bias, we computed the F statistics [38] as:

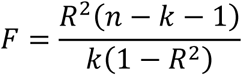

where *k* is the number of SNPs in the instrument and *n* the sample size of the exposure GWAS. *R*^2^ is the total proportion of explained variance for each exposure, calculated by summing all *R*^2^ values of all variants included in the instrument. It can be computed, using the following formula [39]:

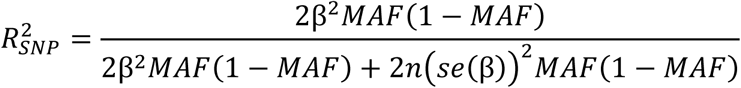

where MAF is the minor allele frequency, *β* is the effect size for a given variant and *n* the sample size of the exposure GWAS. Instruments with F >10 were considered sufficiently strong and retained for analysis.

To reduce computational time in the clumping process, we used a local version of the European population (503 samples) genotypes from the 1000 Genomes Project (phase 3) [40], [41], that excluded variants with minor allele frequency (MAF) less than 1%.

### Methods for causal inference

After the general QC of GWAS, two-sample MR analysis was performed, using “TwoSampleMR” version 0.6.8 package [42], [43] in R environment version 4.4.1. The main MR method we employed to evaluate causality is the “Inverse Variance Weighted” (IVW) method [44], [45], as it is the strongest method with the greatest discovery power. To avoid the risk of overestimating the significance of our findings, we applied a correction for multiple testing within each of the 1490 outcomes – the Benjamini-Hochberg (BH) false discovery rate (FDR) correction [46] – considering a significance level of 5% after correction.

The MR method relies on three important assumptions: (1) IVs are associated with exposure (relevant exposure); (2) IVs are not associated with outcome due to confounding pathways (independence); (3) IVs do not impact the outcome directly, except potentially via exposure (exclusion restriction) [44].

While the first assumption is met by our selection of IV with the clumping method, the other two assumptions cannot be investigated *a priori*. Therefore, in addition of the IVW method, we employed other MR methods as sensitivity analyses: weighted median method [47] and MR-PRESSO (using “MRPRESSO” version 1.0 package on R) [48], [49]. Given the limited power, we considered significant those tests with nominal p-value less than 0.05 (thus not applying multiple testing correction). Of note, while we derivate causal estimates also from the MR Egger test and these are provided on Supplementary Tables, they were not used to define a causal relationship significant, as these estimates have inflated Type 1 error rates [50]. Nevertheless, even if not significant in some cases, the direction of the causal effect was concordant with other MR methods. In addition, we specifically investigated the presence of constant horizontal pleiotropy, heterogeneity, and the influence of a single variant on causal estimates with several statistical tests [50]. In particular, these included the Egger intercept term which assumes all IVs display the same amount of pleiotropy, the Cochran’s Q statistics which assesses heterogeneity across single IV causal estimates [51], and the leave-one-out test – where IVW is repeated removing in turn one IV from the set of IVs. The results were also visualized graphically using scatter plots and leave-one-out plots.

Significant results after IVW were also subjected to bidirectional MR analysis, it means using the outcome as exposure and the exposure as outcome, to confirm the lack of significant reverse causal relationships. This approach ensures that observed associations are not merely due to reverse causation or confounding, thereby strengthening the validity of causal inferences drawn from the study.

We then run replication analyses for all significant findings (those passing IVW and sensitivity analyses), using independent GWAS’ datasets as outcome and exposure, if available (**Supplementary Table 1c**).

### Post-hoc power analysis

For all our significant results we calculated the post-hoc power to compare the power of the main and replication analyses and have further information about the strength of our findings. We used for continuous outcomes the formula [52]:

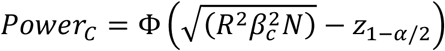

and for binary outcomes:

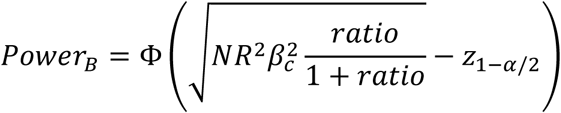

where *β*_*c*_ is the causal effect obtained from main MR analysis, N is the sample size of the outcome’s GWAS, *R*^2^ is the exposure’s explained variance by the selected IVs and *ratio* is calculated by dividing the number of cases by the number of controls of the outcome GWAS. The function Φ is the cumulative distribution function of the standard normal distribution and *z*_1−α/2_ is the quantile of the standard normal distribution corresponding 1 − α/2, for a significance level α = 0.05.

To compute the explained variance *R*^2^, required to calculate the statistical power, we used the formula described in the QC section [39].

### Further investigation of GalNAc-linked utilization

Since in the study by Lopera-Maya et al. [22] the LACTOSECAT pathway was highly associated to a genetic variant in the *ABO* gene, the gene responsible for determining blood group types (**Supplementary Table 1**), we further investigated the significant relationships related to this pathway. A recent study by our collaborators [23] has shown that *ABO*-associated species can also utilize N-acetylgalactosamine (GalNAc) sugar from blood type A individuals who can secret the mucosal A-antigen. We therefore aimed to investigate whether the causal relationships observed in our study might be connected to utilization of GalNAc in blood type A individuals.

Specifically, we assessed the causal relationships between the presence of structural variants (SVs) of *Faecalibacterium prausnitzii*, which contain genes that perform GalNAc degradation activity [53], and the LACTOSECAT pathway and significant outcome results. Of note, this bacteria species was not detected in Lopera-Maya et al. [22], and thus no GWAS was available for our MR analyses. However, the study of Zhernakova et al. [37] identified human single nucleotide polymorphisms (SNPs) that alter the abundance of the GalNAc utilization gene region, and thus these SNPs could be used to investigate the causal effect of *F. prausnitzii*.

We conducted MR analysis using the abundance of the deletion SV (dSV) region (577-579 kb) related to GalNAc activity in *F. prausnitzii* as exposures (**Supplementary Table 1d**), with proteins and lactose-galactose degradation I pathway and significant related phenotypes as outcomes.

We performed clumping of GWAS related to abundance of the dSV with the same criteria used in the main analysis and then we tested the causal relationships with MR method, considering as significant the results with IVW p-value < 0.05.

## Data Availability

No data was generated for this study. We used public data, download links are available in Supplementary Table 1. The scripts used for all the analyses are publicly available in the GitHub repository: https://github.com/Sanna-s-LAB/Mendelian-randomization-Project.git

## Abbreviations

AMD: Age-related macular degeneration
BH: Benjamini-Hochberg
C7: Complement component C7
CD3G: T-cell surface glycoprotein CD3 gamma chain
dSV: deletion structural variant
FDR: False Discovery Rate
GalNAc: N-acetylgalactosamine
GV: Genetic variant
GWAS: Genome-wide association study
HDL: high-density lipoproteins
IV: Instrumental variable
IVW: Inverse Variance Weighted
LD: Linkage disequilibrium
MAF: Minor allele frequency
MR: Mendelian Randomization
MR-PRESSO: MR-Pleiotropy Residual Sum and Outlier
QC: Quality control
SERPINA3: Alpha-1-antichymotrypsin
SFTPD: Pulmonary surfactant-associated protein D
SIGLEC8: Sialic acid-binding Ig-like lectin 8
SNP: Single-nucleotide polymorphism
STROBE: Strengthening the reporting of observational studies in epidemiology
TREM2: Triggering receptor expressed on myeloid cells 2
UKB-PPP: UK Biobank Pharma Proteomic Project
WM: Weighted Median

## Authors contributions

Conceptualization: SS

Project Supervision: DZ, SS

Statistical analyses: FG

Manuscript Draft Preparation: FG, DZ, SS Funding: SS

## Acknowledgements

We thank Daria Zhernakova and Valeria Lo Faro for critical revisions on the manuscript, and Davide Murrau for support and management of our IT infrastructure.

## Ethical Statement

For this study only summarized non-individual data was used.

## Availability of data and materials

No data was generated for this study. We used public data [16], [17], [19], [20], [21], [22], [53], [54], [55], [56], [57], [58], [59], [60], [61], [62], [63], [64], [65], [66], download links are available in **Supplementary Table 1**. The scripts used for all the analyses are publicly available in the GitHub repository: https://github.com/Sanna-s-LAB/Mendelian-randomization-Project.git

## Use of AI statement

We acknowledge the use of artificial intelligence tools, namely ChatGPT 4.0, to improve code efficiency and parallelization.

